# US-Guided Fine-Needle Aspiration of Papillary Thyroid Microcarcinoma in Non-Screening Settings: Diagnostic Errors and Mitigation Strategies

**DOI:** 10.64898/2026.10.06.26364827

**Authors:** Imad A El Hag, Mohamed T Al Shammary, Raghad Tallab, Sharanamma Patil, Abrar Alghamdi

## Abstract

**Background:** Papillary thyroid microcarcinoma (PTMC, ≤1 cm) is increasingly diagnosed, and a subset behaves aggressively. Preoperative ultrasound (US) and fine-needle aspiration (FNA) accuracy in this size range, particularly with multinodular disease, is poorly defined outside East Asian screening cohorts.

**Materials and Methods:** We retrospectively analyzed 180 consecutive PTMC cases (2017– 2026) with preoperative US and FNA and compared them with 191 benign thyroidectomy controls. ROC analysis identified an optimal size cutoff. We compared sensitivity, specificity, PPV, NPV, and accuracy across tumor-size subgroups, before/after 2023, and with/without dominant nodules (DNs) ≥2 cm.

**Results:** ROC analysis identified 5 mm as the optimal cutoff (AUC 0.80). FNA sensitivity was 44.4% overall, falling to 14.1% for tumors <5 mm versus 61.2% for ≥5 mm, with specificity uniformly high (94.8%). US showed a similar pattern (sensitivity 23.4% vs. 64.7%) but lower specificity (69.6%), driven largely by calcification and Hashimoto’s-related hypoechogenicity. Combined FNA+US improved sensitivity to 59.4%. DNs ≥2 cm markedly reduced FNA true-positive rates (67.9% vs. 10.8%; OR 17.4). Excluding DN cases raised FNA sensitivity to 67%, NPV to 84.2%, and accuracy to 85.2%. Post-2023 experience significantly improved all metrics (sensitivity 34.4% to 54.4%).

**Conclusion:** FNA and US sensitivity for PTMC decline sharply below 5 mm and in multinodular glands with dominant nodules, while specificity/PPV remain high. A 5 mm threshold, operator experience, and selective avoidance of FNA in dominant-nodule–bearing multinodular goiters (favoring direct surgical referral) can meaningfully improve diagnostic performance and guide management.

## Introduction

Papillary thyroid microcarcinoma (PTMC), defined as a tumor measuring 1 cm or less, has long been regarded as an indolent subtype of papillary thyroid carcinoma ^1^. These tumors are often discovered incidentally in thyroids that are removed for benign conditions or during postmortem examinations ^2, 3, 4^. Most cases tend to go unnoticed throughout a patient’s lifetime. However, with recent advances in imaging technology and the increasing use of ultrasound-guided fine-needle aspiration (US-FNA) for monitoring thyroid nodules, there has been a significant increase in preoperative diagnoses ^5^. Today, more than 50% of newly diagnosed papillary thyroid carcinomas (PTCs) worldwide are 1 cm or smaller.

The aggressive forms of this non-incidental subgroup behave clinically similarly to, and in some cases worse than, their counterparts that exceed 1 cm in size ^6^. These aggressive tumors have documented instances of recurrence, locoregional and distant metastases, and disease-related mortality. As a result, the 2022 WHO Classification of Thyroid Neoplasms no longer recognizes PTMC as a distinct subtype ^7^. Instead, it categorizes these tumors as early-stage papillary thyroid carcinoma (pT1a), subjecting them to the same risk stratification and management guidelines as tumors larger than 1 cm. Tumors that exhibit aggressive behavior typically exceed 5 mm in size, have a higher prevalence of BRAF mutations, and display specific morphological subtypes, including diffuse sclerosing, tall cell, oncocytic, hobnail, columnar, and solid variants ^8, 9, 10, 11^.

Because PTMCs represent a significant portion of the global burden of PTC and may behave similarly to larger tumors, accurate preoperative diagnosis is essential for guiding treatment decisions, whether that involves immediate surgery or active surveillance. Yang et al. first reported the effectiveness of US-FNA in diagnosing PTMC. In a non-case-control study of 39 patients, they showed the procedure could detect tumors as small as 2 mm. Later case-control studies have confirmed that US-FNA can effectively diagnose sub-centimeter thyroid carcinomas. These studies reported sensitivities of 84.5% and specificities of 96.8%, figures comparable to those for tumors larger than 1 cm, which had sensitivities of 96.7% and specificities of 95.7% ^13^. Overall, the average sensitivity and specificity of US-FNA are 72% and 83%, respectively, indicating no significant difference in diagnostic performance between tumors below and above 1 cm (14).

Because of its superficial location, the thyroid gland is primarily evaluated by ultrasound. This method is low-cost, widely available, and quick, making it the main diagnostic tool for thyroid diseases. Ultrasound is often used in conjunction with FNA cytology during the preoperative assessment of thyroid nodules. Additionally, it can be employed independently in mass screening programs for carcinomas that measure 1 cm or larger, as well as for patients undergoing active surveillance ^3, 15, 16^. The combination of ultrasound and FNA has been found to be more effective than either method used alone for detecting tumors 1 cm or smaller.

Several factors affect the diagnostic accuracy of US-FNA for small nodules, including operator experience, tumor size, and whether the nodules are solitary or multiple. Research specifically examining the impact of tumor size on FNA accuracy in PTMC has shown no significant difference in outcomes between tumors smaller or larger than 5 mm ^18,19^. However, the potential influence of a dominant nodule within a multinodular thyroid gland has not yet been thoroughly investigated. Most of the published evidence supporting the effectiveness of ultrasound-guided FNA in diagnosing small papillary carcinomas comes from East Asia. In this region, high-volume screening programs have typically identified normal glands with small, isolated nodules, leading to significant institutional expertise ^3, 15, 18, 19^. In contrast, experience in the Middle East and Western countries is limited and is primarily derived from clinical contexts that do not involve screening.

Despite the growing literature on this topic, data on a clinically meaningful threshold for tumor size remain limited. This information is crucial, as tumor size affects biological behavior and guides management decisions. This study aims to identify such a threshold in a general hospital setting, establish a practical benchmark for reliable diagnostic performance, and highlight avoidable sources of diagnostic error. Given the two main management strategies for PTMC, immediate surgical resection versus active surveillance, the diagnostic accuracy of FNA and ultrasound, both individually and in combination, will be evaluated within our institution and compared with published data.

## Materials and Methods

### The study population

This study encompasses all consecutive cases of papillary thyroid microcarcinoma (PTMC) measuring 10 mm or less that were diagnosed at our institution between January 2017 and March 2026. The research received approval from the Institutional Review Board (H-01-R-069), and informed consent was waived due to its retrospective nature. For every case included, preoperative ultrasound (US) and fine-needle aspiration (FNA) cytology data were available. Additionally, a control group matched by age and sex, consisting of patients who underwent thyroidectomy for benign conditions, was selected from the same period for comparison.

Sociodemographic variables, including age at diagnosis and sex, were recorded prospectively at the time of initial evaluation. Tumor size was determined by integrating gross pathological measurement with histological assessment; only tumors with a maximum diameter of 10 mm or less on final pathological examination were eligible for inclusion. In multifocal disease, the largest (dominant) focus was used for all size-based analyses.

### FNA performance and reporting

All thyroid fine needle aspirations (FNA) are performed under ultrasound guidance by an interventional radiologist using 25-gauge needles. On average, four passes are made, with the specimens directly deposited onto four slides. Direct smears are prepared, with two air-dried for Diff-Quick staining and the other two alcohol-fixed for Pap staining. The adequacy evaluation is carried out on-site. All thyroid FNAs are reported using the six-tiered Bethesda System for Reporting Thyroid Cytology ^20^. Reporting of thyroid FNA is limited to a few pathologists who are supervised by a cytopathologist with an interest in the field. BSRTC categories IV, V, and VI are considered positive because they prompt thyroidectomy.

### Ultrasound performance and reporting

The ultrasound examination typically precedes the FNA procedure, with ultrasound findings, among other factors, used to determine whether to proceed with a US-FNA. Ultrasound findings are reported according to the American College of Radiology Thyroid Imaging, Reporting and Data System (ACR-TIRADS), which is based on tumor composition, echogenicity, shape, margins, and the presence of calcifications ^21^. It is a five-tiered system, with classes 3, 4, and 5 carrying the highest risk of malignancy and requiring subsequent FNA. Gland volume, number of nodules, and size of the dominant nodules are determined during the first ultrasound examination. Both the ultrasound and gross findings are used to determine the size of the dominant nodule.

### Receiver operating curve (ROC) in tumor classification

The receiver operating characteristic (ROC) curve and area under the curve (AUC) analyses function as tools for binary classification. They help identify the tumor size at which US-guided FNA achieves optimal sensitivity and specificity. In this analysis, tumor size is plotted against the results of the fine-needle aspiration.

### Sensitivity and specificity for FNA and ultrasound

Sensitivity and specificity were assessed across three size-defined subgroups: tumors measuring 1–10 mm (Group A), tumors that met or exceeded the cutoff size derived from the ROC analysis (Group B), and tumors that fell below this cutoff (Group C). The clinical implications of these diagnostic parameters for patient management are also discussed. We evaluated the sensitivity, specificity, and diagnostic performance of FNA, ultrasound, and their combination across these groups. To establish a reference standard, we formed a control group consisting of consecutive cases with histologically confirmed benign diagnoses obtained during the same study period. All members of this group had undergone preoperative ultrasound and FNA. For classification purposes, BSRTC categories II and III and ACR TI-RADS categories 1, 2, and 3 were considered true negatives (TN) when subsequent surgical pathology confirmed a benign lesion or benign neoplasm. They were classified as false negatives (FN) when the final histological diagnosis was PTMC. Conversely, BSRTC categories IV, V, and VI and TI-RADS categories 4 and 5 were classified as true positives (TP) when the final diagnosis was PTMC, and as false positives (FP) when pathology revealed a benign lesion or benign neoplasm.

### False-negative and false-positive rates and causes in thyroid Ultrasound and FNA

The main causes of FN and FP results in diagnosing PTMC are examined, including cytological oversights and interpretation errors.

### Role of operator experience in US-guided FNA performance for PTMC

The effect of operator expertise on preoperative diagnostic performance is assessed by comparing sensitivity and specificity before and after the significant increase in PTMC diagnoses noted in 2023 ^22^.

### Impact of case selection on US-guided FNA performance

The effect of retrospectively excluding cases involving multinodular glands with dominant nodules (DNs) on the diagnostic performance of US-guided FNA is examined. Cases featuring enlarged multinodular glands harboring DNs were excluded; sensitivity and specificity were then determined in the remaining cases without DNs.

### Statistical analysis

All statistical analyses were performed using SPSS version 29.0.2.0. Differences between two proportions or means were evaluated using 95% confidence intervals (CIs), with non-overlapping CIs indicating statistical significance at *p* < .05.

## Results

### Study population

This study examined 180 cases of PTMC identified among 1,549 thyroidectomies performed between January 2017 and March 2026. The patients ranged in age from 20 to 89 years, with a mean age of 43 ± 12 years. The PTMC group was predominantly female, with 150 females and 30 males, for a female-to-male ratio of 5:1. A control group of 191 cases of benign thyroidectomy was selected from the same surgical pool and matched to the PTMC cohort by age and sex. This control group also had a majority of females, comprising 163 women and 28 men, giving a female-to-male ratio of approximately 6:1. The ages of the control group ranged from 15 to 80 years, with a mean age of 44 ± 12 years.

### ROC analysis

The ROC analysis of FNA results compared to tumor size produced an AUC of 0.80 (95% confidence interval: 0.73–0.86), indicating excellent model performance that surpasses random guessing. The highest sensitivity and specificity were 82% and 64%, respectively, for tumors measuring 5 mm, the model’s defined cut-off size (Figure 1).

**Figure 1.**
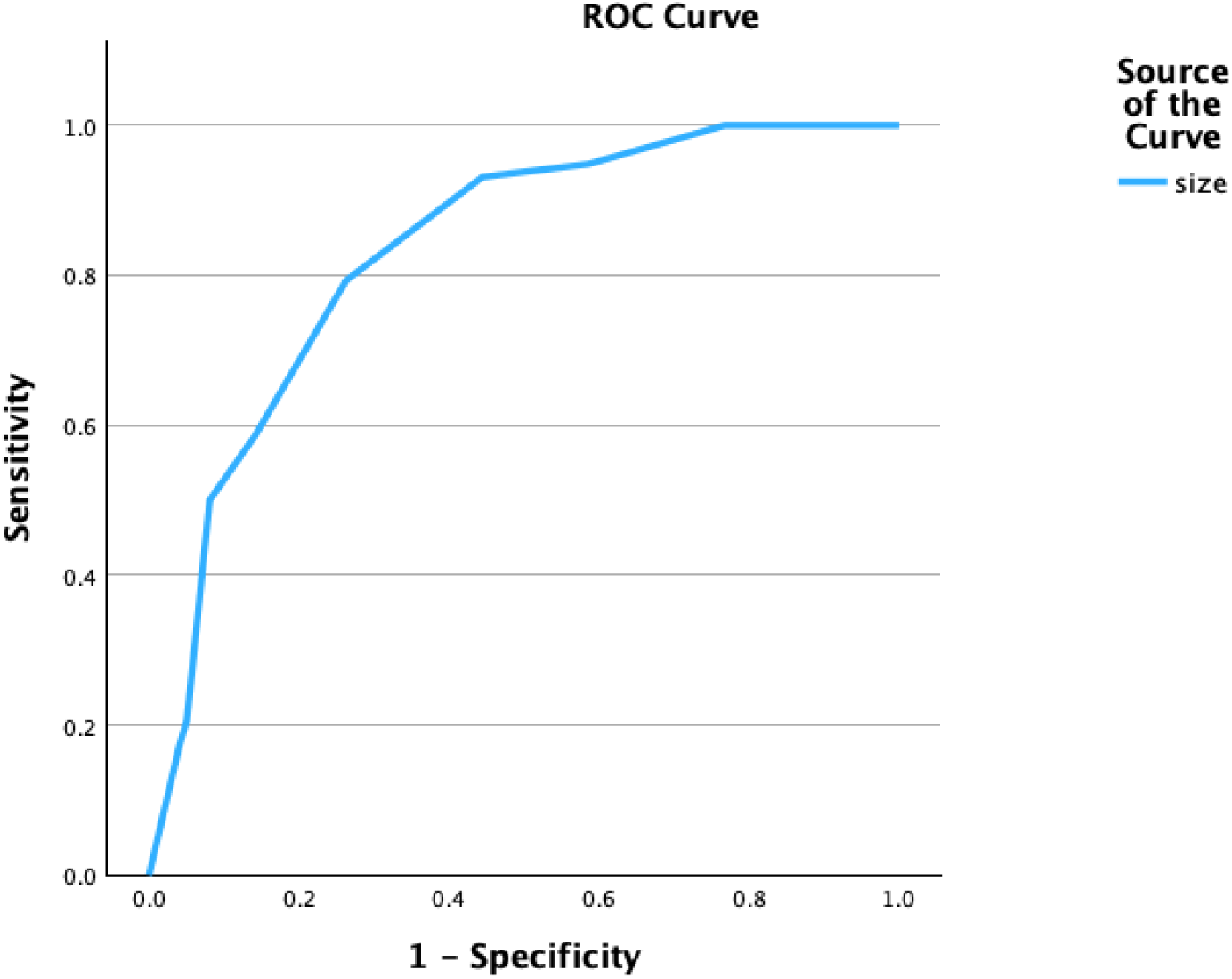
ROC analysis curve for tumor size versus FNA specificity and sensitivity.

### Accuracy of FNA and US, separate and combined

Figures 2, 3 & 4 summarize the diagnostic performance of FNA and ultrasound, separately and combined, for detecting PTMC, including sensitivity, specificity, positive predictive value (PPV), negative predictive value (NPV), and accuracy.

**Figure 2.**
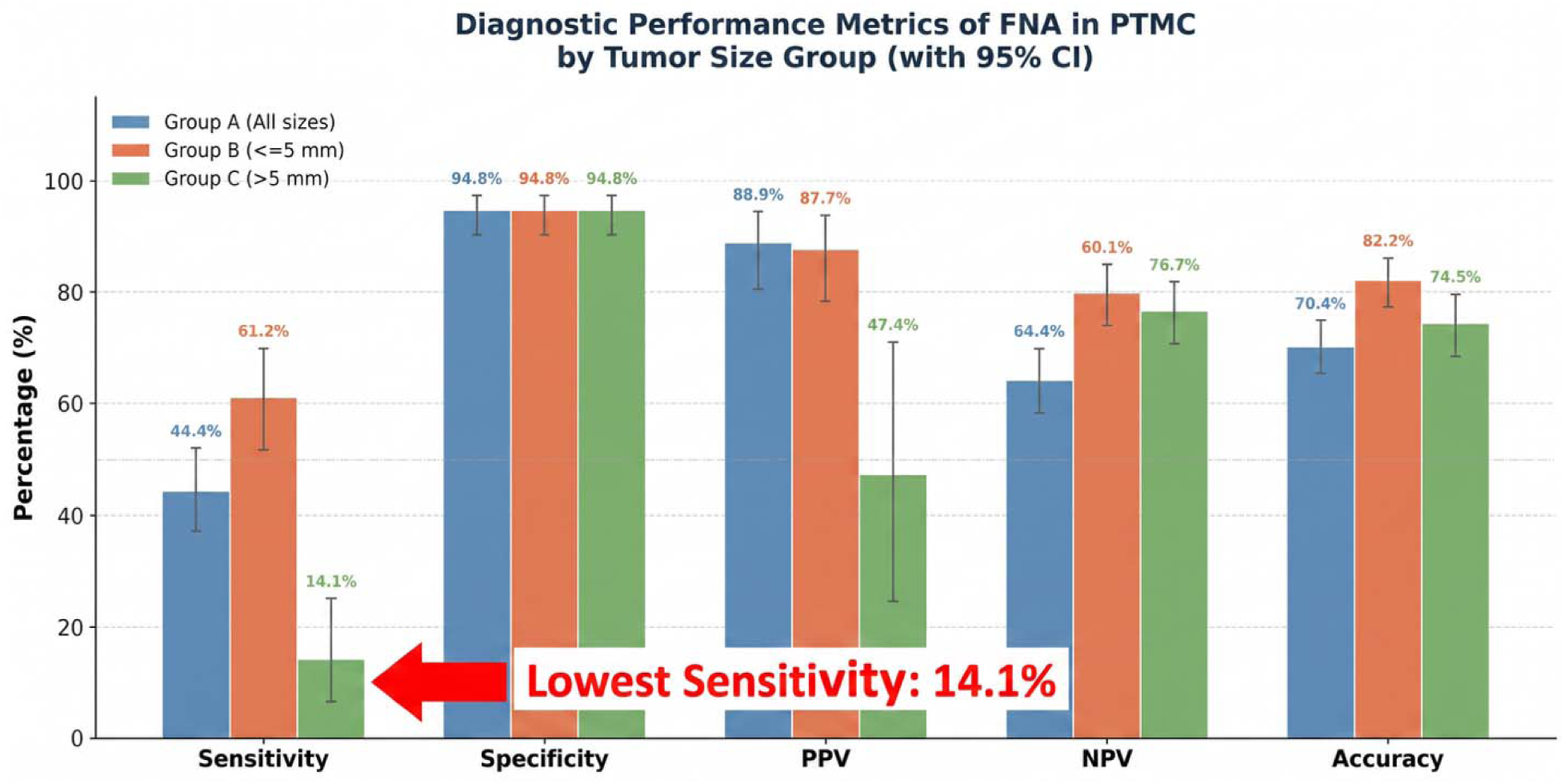
Diagnostic performance metrics of FNA in PTMC by tumor size group (with 95%CI).

FNA results (see Figure 2) demonstrate the procedure’s effectiveness across three PTMC tumor size categories. FNA correctly identified 80 of 180 positive cases as malignant in group A; 71 of 116 in group B; and 9 of 64 in group C. The control group, consisting of 191 benign cases, had 10 false positives and 181 true negatives.

The test consistently achieved very high specificity (94.8%) for diagnosing PTMC across all groups. However, sensitivity varied widely, with an overall sensitivity of 44.4%, performing best in the ≥5 mm group (61.2%) and worst in the >5 mm group (14.1%). Group B (≥5 mm) showed the highest overall diagnostic accuracy (82.1%). Conversely, Group C, which included tumors smaller than 5 mm, performed poorly: FNA detected only 1 of 7 malignant cases, reducing the PPV to 47.4%. Overall, FNA performance differed significantly by tumor size (χ² = 57.08; p < .001). Notably, the smallest PTMC diagnosed as cytologically malignant (BSRTC category IV) in our series was 3 mm.

The US results (see Figure 3) show the procedure’s effectiveness across three PTMC tumor size groups. In group A, US correctly identified 90 of 180 positive cases as malignant; in group B, 75 of 116; and in group C, 15 of 64. The control group, with 191 benign cases, had 10 false positives and 181 true negatives.

**Figure 3.**
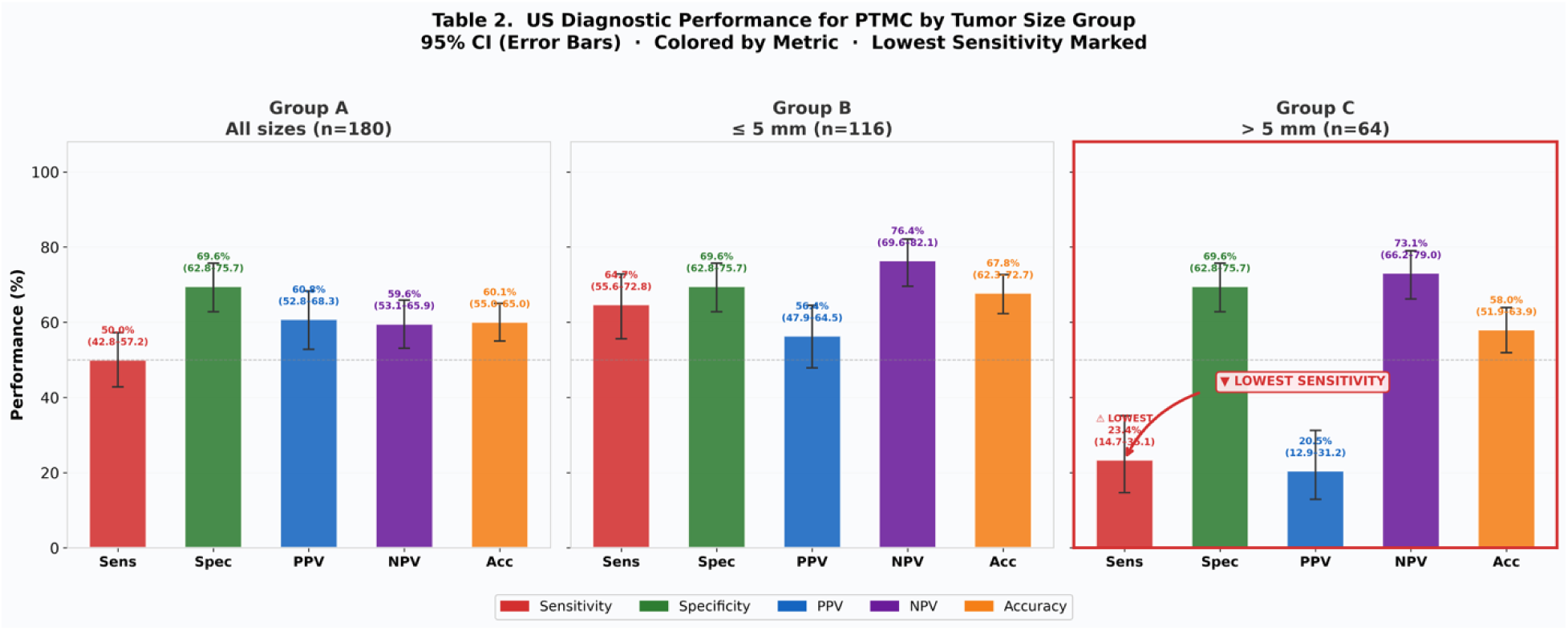
Ultrasound diagnostic performance for PTMC by tumor size group (95% CI)

A similar pattern was observed in the US cohort’s sensitivity measurements (see Figure 4). Sensitivity declines sharply from 64.7% for tumors larger than 5 mm to 23.4% for tumors 5 mm or smaller, a threefold reduction. Specificity remains steady at 69.6% across all groups. The PPV drops to 20.5% in Group C, indicating that only 1 in 5 nodules labeled as ‘malignant’ by US are truly malignant. NPV rises from 59.6% in Group A to 73.1% in Group C, reflecting the lower malignancy prevalence in the ≤5 mm subgroup. Overall accuracy remains low, ranging from 58% to 68% across all groups. Differences in diagnostic performance among Groups A, B, and C were highly statistically significant (χ² = 28; p < .001). Group B demonstrated significantly superior malignancy detection compared with both Groups A and C, while Group C had the lowest overall detection rate.

**Figure 4.**
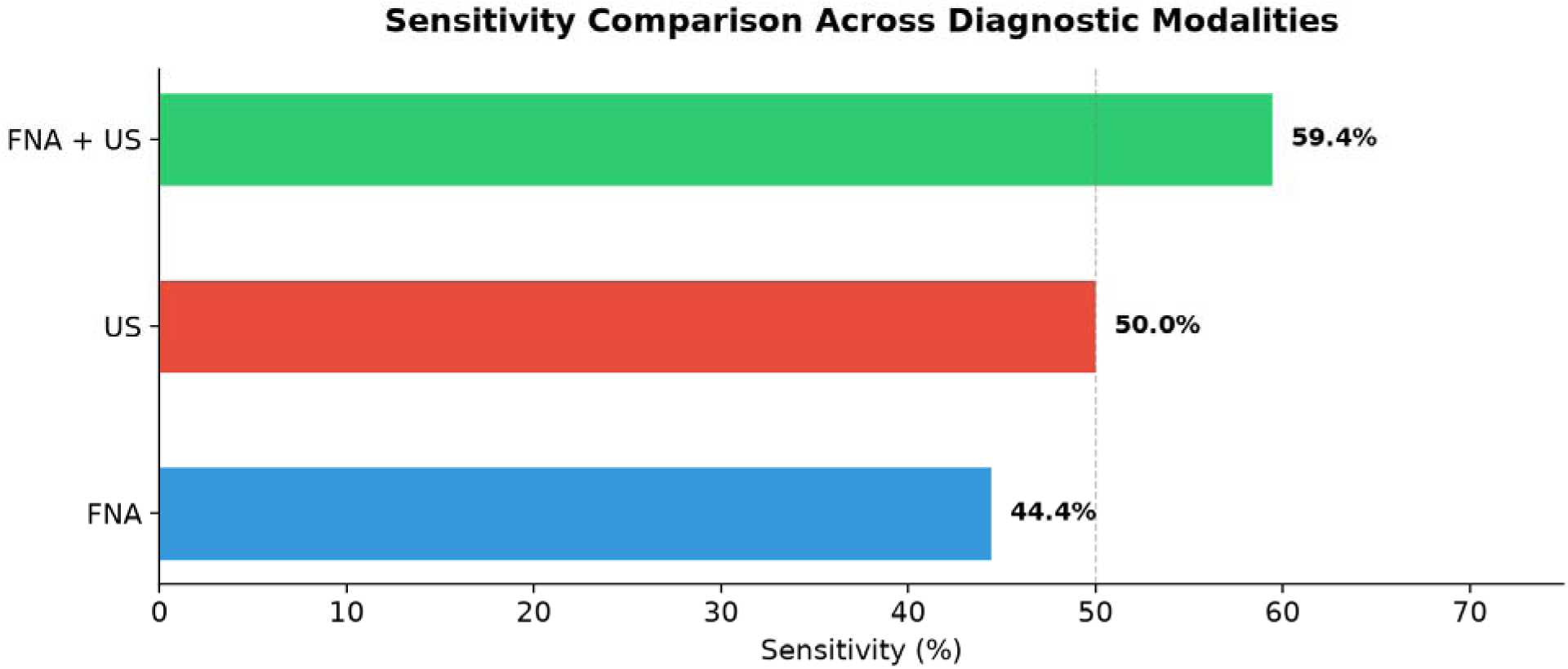
Sensitivity comparison across the three diagnostic modalities FAN, US, and FNA+US)

A combined FNA & US identified 107 of 180 cases as true positive, while the control group of 191 had 64 positive. Figure 4 compares FNA, US, and their combination. All three show a typical sensitivity–specificity trade-off: FNA alone exhibits the highest specificity (94.8%) and PPV (88.9%), but the lowest sensitivity (44.4%), missing over half of PTMC cases. Ultrasound alone provides moderate sensitivity (50.0%) with notably lower specificity (69.6%). Combining FNA + US yields the highest sensitivity (59.4%), a 15% increase over FNA alone and a 9.4% increase over US alone, though it reduces specificity to 66.5%. The Cochran Q test reveals a significant difference among the three diagnostic methods (Q = 22.82; p < .001).

From a clinical Bayesian perspective, the high PPV of FNA (88.9%) indicates that when FNA returns a malignant result, there is a very high probability of true disease, making FNA a powerful rule-in test. Conversely, the NPV of all three modalities remains moderate (59.6%–64.4%), meaning that a negative result from any single modality cannot reliably exclude PTMC.

### False negative and false positive rates in FNA and their causes

Table 1 shows the false-negative rates of FNA in diagnosing PTMC along with their underlying causes. In Group A, which includes all tumors regardless of size, the overall FN rate is 55.6%. In the group of tumors smaller than 5 mm, the TP rate is very low (9 out of 64 cases, or 14.1%). In contrast, tumors 5 mm or larger have a TP rate of 61.2%, while tumors between 1 and 10 mm have a TP rate of 44.4%. This suggests tumor size strongly predicts a positive FNA diagnosis. Furthermore, group B had a 9.64-fold higher odds of a positive FNA result (OR 9.64, 95% CI 4.34-21.4; P <0.001). The FP rate is as low as anticipated, with 10 cases out of 191 in the control group, resulting in a rate of 5.2%. The primary reason for this rate is that in 90% of the false-positive cases, a cellular hyperplastic nodule within a goiter was incorrectly classified as category IV BSRTC.

**Table 1.** False negative rate in FNA across different tumor sizes.

### The significance of dominant nodules (DNs) on FNA results

To explore factors contributing to FN rates beyond small tumor size, we investigated whether a multinodular goiter with a dominant nodule larger than 2.0 cm, in conjunction with PTMC, affected FNA diagnostic performance. Table 2 depicts the findings.

**Table 2.** Impact of DNs included vs. excluded on FNA results.

We found that the FNA true-positive rate was 67.9% in cases without DNs but dropped significantly to 10.8% when DNs were present. This resulted in an odds ratio of 17.4 (95% CI: 7.57–39.92), indicating that the absence of DNs was associated with a 17-fold higher likelihood of receiving a positive FNA result. This difference was highly statistically significant (p < 0.001).

These results suggest that the presence of DNs is associated with an increased likelihood of false-negative FNA results.

Notably, the primary PTMC associated with diffuse nodular disease was significantly smaller, measuring 4.76 ± 2.2 mm, compared with those without DNs, which averaged 6.03 ± 2.6 mm (F = 11.5, p < 0.001). This suggests that small tumors, particularly those under 5 mm, are more commonly found in cases of multinodular thyroid disease with DNs. The coexistence of small tumor size and DNs creates a combined effect that further complicates accurate FNA targeting.

In summary, the main factors contributing to the anticipated FN results in this study were small tumor size and the presence of DNs, with substantial overlap between these two factors. Additional contributing factors included interpretive challenges stemming from specimen heterogeneity and poor cellularity, which led to classifications as Atypical Undetermined Significance (Bethesda Category III). Importantly, none of the false-negative cases in this series were due to interpretive errors or inaccurate smear reporting.

### False negative and false positive rates in ultrasonographic evaluation of thyroid and their causes

Table 3 provides the FN rates for US examinations. The overall FN rate for Group A was 50%, which increased significantly to 76.2% for tumors smaller than 5 mm. Additionally, compared with Group B, Group C had significantly lower odds of a positive US result (OR 0.17, 95% CI 0.089-0.33; P <0.0001).

**Table 3.** False negative rate in US across different tumor sizes.

Group C had approximately six times lower odds of receiving a positive US result than Group B (p < 0.001). FP diagnoses in thyroid ultrasounds are becoming a significant clinical concern. Among TIRADS classes 4 and 5, 58 out of 191 negative cases were mistakenly classified as positive, resulting in an overall false-positive rate of 30.4%,

### The significance of DNs

Table 4 highlights the impact of DNs on US performance. The rate of TP ultrasound results significantly decreased from 66.0% in the absence of DNs to 27.0% when DNs were present. This corresponds to a 5.25-fold increase in the odds of obtaining a positive ultrasound result when DNs are not present (p < 0.001). These findings indicate that DNs impair US detection of PTMCs. Additionally, DNs-associated PTMCs tend to be smaller, further complicating the diagnostic challenge, similar to the effects previously described for FNA performance.

**Table 4.** Impact of DNs included vs. excluded on US results.

### Sources of false-positive diagnoses

Table 5 summarizes the main sonographic features that contribute to false-positive diagnoses. These include interpretation errors related to calcification, hypoechoic nodules, especially those associated with Hashimoto’s thyroiditis, taller-than-wide nodules, and granulomas resulting from prior fine-needle aspiration. While calcification is distributed evenly between cases of Hashimoto’s thyroiditis (HT) and non-HT cases, hypoechogenicity occurs significantly more frequently in HT cases (51%) compared to non-HT cases (24.6%) [X = 12.2; P < 0.001].

**Table 5.** Causes and rates of US false positives.

Statistical analysis of these features reveals three distinct behavioral patterns. Calcification is statistically associated with FP diagnoses in benign nodules, whereas hypoechoic texture shows a more balanced distribution across both FP and FN outcomes, suggesting it has less diagnostic significance than calcification. However, when both features are present simultaneously, as in 15 cases in this study, their combined effect is significantly stronger. The additive odds ratio for this combination is approximately 14.4, indicating that a patient with both characteristics is roughly 14 times more likely to be a false positive than a true negative.

Because of the small sample size, nodules that are taller than wide and granulomas from prior fine-needle aspirations could not be evaluated statistically. Nevertheless, their potential to cause false positives should not be overlooked.

### Impact of experience on the performance of US-guided FNA for PTMC

Figure 5 summarizes the results. In 2023, we observed a significant increase in PTMC cases at our institution. As a result, we assessed the sensitivity, specificity, PPV, NPV, and accuracy of US-guided FNA both before and after 2023.

**Figure 5.**
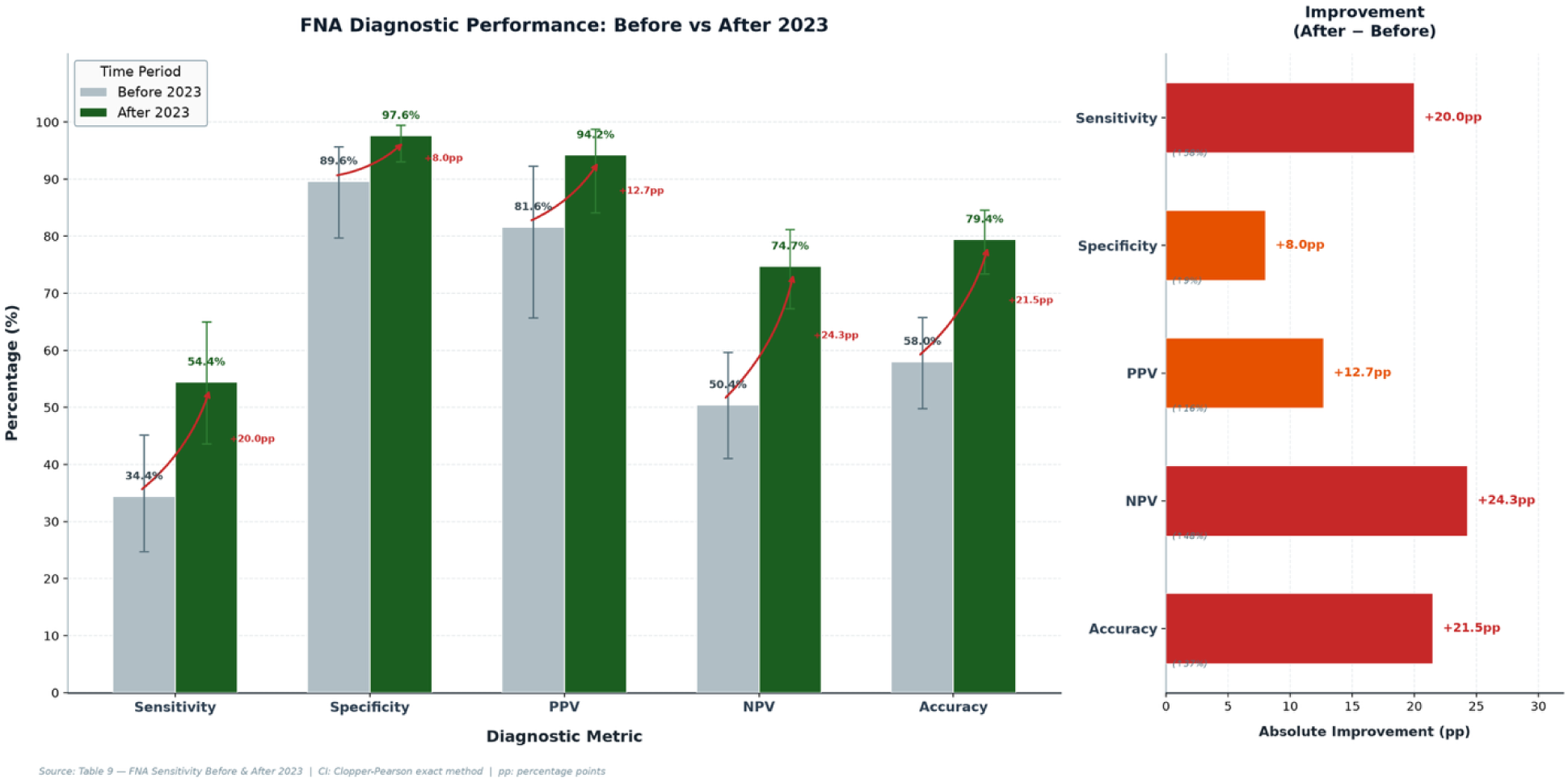
FNA diagnostic performance before and after 2023

Before 2023, FNA correctly identified 31 malignant cases out of 90, with a control group of 67 benign cases that had 7 false positives. After 2023, the procedure correctly identified 49 malignant cases out of 90, while a control group of 124 benign cases had 3 false positives.

The analysis shows substantial improvement across all five key diagnostic metrics in the post-2023 period, with particularly notable gains in negative predictive value and sensitivity. Sensitivity rose from 34.4% to 54.4%. Specificity rose from 89.6% to 97.6%. PPV rose from 81.6% to 94.2%. NPV rose from 50.4% to 74.7%. Accuracy rose from 58.0% to 79.4%. The most clinically significant finding is the NPV increase from 50.4% to 74.7%, which transforms the reliability of a negative FNA result from essentially a coin flip to approximately three-quarters reliable, a potentially practice-changing shift in clinical decision-making for papillary thyroid microcarcinoma (PTMC). The most clinically meaningful gain was in sensitivity, as missing a malignancy poses the greatest risk to patients (χ² = 7.29; p < 0.01).

The average tumor size before and after 2023 remained relatively constant, measuring 5.3 ± 2.4 mm and 5.5 ± 2.8 mm, respectively (F = 0.28; p = 0.59). Additionally, the proportion of multinodular glands with DNs remained stable over both periods, at 43% and 42%, respectively (χ² = 0.016; p = 0.52). These findings suggest that changes in tumor size and the presence of DNs did not affect the procedure’s performance.

One remaining concern is that even after 2023, the sensitivity of 54.4% is still modest for a cancer diagnostic tool, indicating that nearly half of malignant cases may still receive a benign FNA result.

### Impact of case selection on the performance of US-guided FA

Figure 6 examines the five performance metrics for Fine Needle Aspiration (FNA) before and after removing cases with DNs. With DNs included, FNA correctly identified 80 of 180 (44.4%) cases as malignant. After excluding cases with DNs, it identified 72 of 106 (67%) cases as malignant. The control group consisted of 191 benign cases, and 10 false positives were observed regardless of DN presence. The most notable improvement is in sensitivity. When DNs are present, FNA detects over half of malignancies, with a sensitivity of just 44.4%. Excluding DNs raises the sensitivity by 23.5 percentage points to 67%. The non-overlapping 95% confidence intervals (37.2%–51.7% vs. 59.0%–76.8%) confirm this is a statistically significant improvement. Specificity stays high at 94.8% in both groups because excluding DNs does not affect FNA’s ability to correctly identify benign nodules—the control group (true benign nodules) and the correctly identified benign cases (181 TN, 10 FP) remain unchanged. DNs influence only the malignant cases in the analysis. Another key improvement is in NPV, which increases from 64.4% with DNs to 84.2% without DNs, a 31% relative increase. When DNs are excluded, a “benign” FNA result is far more reliable. Previously, a benign result had a 64% chance of being correct; now, it’s 84%. The 95% CIs do not overlap (58.8%–70.0% vs. 79.3%–89.1%), confirming significance. Overall accuracy also improves significantly, rising from 70.4% to 85.2%, nearly a 15 percentage point increase driven mainly by fewer false negatives.

**Figure 6.**
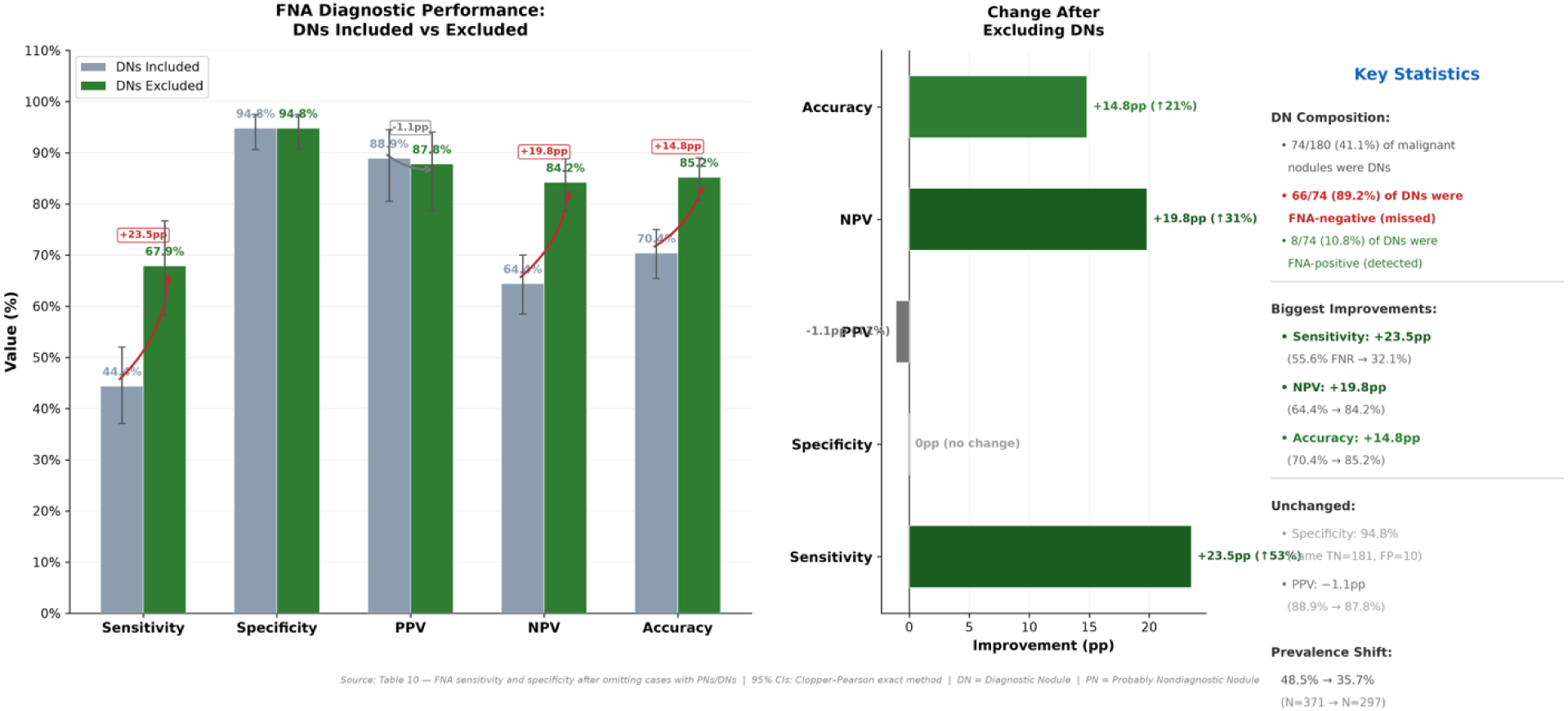
FNA diagnostic performance: DNs included vs. excluded.

## Discussion

The adoption of ultrasound-guided FNA as a preoperative diagnostic tool has fundamentally transformed thyroid nodule management, reducing unnecessary surgeries for benign disease while appropriately directing surgical intervention toward confirmed malignancy. Surgical intervention rates are now predictably stratified: 12.9% for benign, 24.3% for indeterminate, and 64.9% for malignant cytologic categories ^23^. The malignancy yield among surgically resected nodules has risen from 18% before FNA to 40% after its introduction ^24^. These benchmarks confirm that FNA meaningfully improves triage efficiency.

However, the diagnostic landscape for papillary thyroid microcarcinoma (PTMC), tumors measuring less than 1 cm, remains substantially more challenging. Over the past three decades, the global incidence of these small tumors has risen markedly, driven by advances in high-resolution ultrasonography, the proliferation of screening programs, and the expanded use of FNA to monitor incidentally discovered nodules ^5^. Environmental contributors, including radiation exposure, iodine deficiency, and evolving lifestyle patterns, may further augment this trend. Critically, a subset of PTMC exhibits aggressive behavior, local recurrence, distant metastasis, and progression to high-grade carcinoma with poor outcomes ^25^, and currently accounts for up to 30.5% of the thyroid cancer surgeries at our institution, with 47% of these cases deemed clinically significant ^22^.

It is within this context that we undertook a systematic evaluation of FNA diagnostic performance across three complementary analytical frameworks: (1) tumor size stratification (groups A, B, and C), temporal comparison before versus after 2023 (Figure 5), and the impact of excluding multinodular goiter with dominant nodules of more than 2,0 cm (Figure 6). Together, these analyses reveal a coherent, if sobering, picture of FNA’s capabilities and limitations in the PTMC setting.

In our study, the overall sensitivity of US-FNA for tumors measuring 1–10 mm was notably low (44.4%). This is significantly lower than sensitivity levels reported in the literature, which typically range from the seventies to the nineties and appear largely independent of tumor size, whether within the 1–10 mm range or exceeding 1 cm.

Interestingly, tumor size significantly affected sensitivity in our cohort. Within the PTMC spectrum, sensitivity is inversely related to tumor size. Sensitivity decreased sharply to 14.1% for tumors smaller than 5 mm, but increased substantially to 61.2% for tumors measuring 5 mm or larger. Receiver operating characteristic (ROC) analysis also supported this size-dependent trend, identifying 5 mm as a clinically relevant threshold, above which sensitivity can reach as high as 82%. Across all three groups, specificity remained uniformly high at 94.8%, confirming that FNA’s ability to correctly identify benign nodules is unaffected by tumor size.

The lower diagnostic accuracy in our study than in earlier reports likely reflects differences in patient characteristics and case selection. Most prior studies primarily included patients with a single nodule. For example, one study encompassed only 25 cases with two nodules (3%) out of 822 patients, with the vast majority presenting with a solitary nodule ^13^. Another study enrolled 363 patients with single nodules, along with 139 patients (27%) who had primarily two nodules ^19^. In contrast, our cohort included a significantly higher proportion of patients with multiple nodules, with 74 out of 180 patients (41%) having between 2 and 9 nodules.

Multinodularity, especially when accompanied by a benign dominant nodule larger than 2 cm, is associated with a remarkably high FN rate of 89%. In contrast, the absence of a dominant nodule is linked to a 17-fold increase in the likelihood of a positive FNA result, underscoring its significant impact on diagnostic outcomes.

Our findings suggest that two primary factors contribute to cytological misses in this context: tumor size smaller than 5 mm and multinodularity when a dominant nodule measures 2 cm or greater. These factors appear to drive the elevated false-negative rate observed in our study. The reasons include difficulty with needle placement and cytological interpretation challenges arising from smear heterogeneity, where significant cells are diluted among benign features in the colloid.

In our study, we found that papillary thyroid microcarcinomas (PTMCs) associated with multinodularity and dominant nodules were typically quite small, averaging just 4.5 mm. This size is notably smaller than the 6 mm average seen in patients without dominant nodules. Tumors of this size are generally considered indolent and are often discovered incidentally, which de-emphasizes the diagnostic challenge they present. This size difference highlights the critical importance of careful fine-needle aspiration (FNA) targeting in multinodular goiter, as the risk of false-negative results increases significantly.

The challenge is further complicated by the dominant nodules themselves, which can compromise diagnostic accuracy. Multiple studies have shown that FNA of thyroid nodules larger than 4 cm tends to be less reliable, resulting in a high rate of false negatives ^26,27,28,29^. This is due to sample heterogeneity and the incomplete sampling of complex architectural lesions. In our cohort, where dominant nodules often exceeded this size, this issue likely contributed to the increased rate of cytological misses.

Excluding multinodular cases from our analysis significantly improved FNA sensitivity from 44.5% to 67.9%, NPV from 46.4% to 84.2%, and overall accuracy from 70.5% to 85.2%. Therefore, we suggest exempting multinodular goiter cases from FNA. Including these cases inflates the false-negative rate and leads to unnecessary procedural costs. Given that large nodules within a multinodular goiter carry a notably lower risk of malignancy, these patients should proceed directly to surgery, bypassing FNA entirely.

The false-positive (FP) rate in thyroid FNA cytology is generally low, reflecting the procedure’s high specificity. In this cohort, the FP rate was 5.2% (10/191 control cases), consistent with expected benchmarks. The majority of these false positives (90%) arose from cellular hyperplastic nodules within a goiter being misclassified as Bethesda category IV. A recent study at our institution addressed this limitation by emphasizing detailed nuclear feature assessment and refining risk stratification within category IV into low- and high-risk subgroups, an approach that substantially reduced the false-negative rate in prospective validation ^30^.

The considerations that apply to FNA also apply to the ultrasonographic examinations used to detect PTMCs in our study. Overall sensitivity is low (50%). For tumors smaller than 5 mm, the sensitivity is even lower, at 23.4%. However, sensitivity improves for tumors that are 5 mm or larger, reaching 64.7%. As a result, the false-negative rate is high, especially for small tumors (less than 5 mm) and for cases with multinodular glands containing suspicious nodules (DNs), at 76.2% and 73%, respectively.

Unlike FNA, ultrasound exhibits low specificity, with a high false-positive rate of 30.4%. This is due to factors such as misinterpretation of calcifications, hypoechoic nodules seen in Hashimoto’s thyroiditis, tumor orientation (taller than wide), and repeat FNAs. The false-positive causes we identified align with those recently reported in other studies ^31^.

We examined the effects of combining FNA with US compared to using each test separately. The combined approach significantly outperforms either test alone, while FNA and US individually do not differ significantly. Notably, compared with FNA alone, the combined FNA and US method detected 28 more cases, resulting in a 15.6% increase in the positive detection rate. This benefit from combining both tests aligns with established expectations. Combined FNA and US has been shown to improve detection of malignant thyroid nodules, particularly smaller tumors under 1 cm. Specifically, the sensitivity was 93.3% for FNA and 86.6% for US, both reaching 100% when used together.

The key difference between our findings and those of others lies in case selection. Previous studies averaged one nodule per case, whereas we encountered many cases with multiple nodules, including large benign ones over 2 cm.

We also investigated how accumulated experience affects procedural performance and whether it can address issues related to small tumor size and multi-nodularity. We assessed the sensitivity and specificity of FNA, calculating performance metrics over two periods: early (2017-2022) and late (2023-2026). The high incidence of PTMC observed in 2023 serves as the dividing point between these two periods. Two confounding factors—tumor size and the presence of DNs— remained constant throughout both periods. We found that FNA sensitivity increased significantly over time, rising from 44.4% to 67.9%, a gain of 23.5%. This improvement indicates a significant enhancement in the ability to detect these tumors, despite consistent tumor size and nodularity throughout the study period. These findings emphasize the importance of gaining experience in this field.

This study has several limitations. First, subtle ambiguities in cytological classification, combined with shifts in disease prevalence over time, may have influenced the calculated performance metrics—particularly in the analysis of temporal trends. Second, the observed improvement in performance after 2023 cannot be attributed to FNA alone, as it may also reflect changes in patient demographics, nodule characteristics, or concurrent advances in imaging technology. Third, excluding diagnostic nodules (DNs) after classification introduces selection bias: the resulting FNA performance estimates represent a hypothetical best-case scenario rather than the diagnostic reality encountered in prospective clinical practice.

In summary, US-guided FNA has substantially improved the preoperative management of thyroid nodules by reducing unnecessary surgeries for benign cases and guiding malignant cases toward surgical intervention. However, its diagnostic accuracy for PTMCs is strongly influenced by two factors: tumors smaller than 5 mm and multinodularity, particularly when a dominant nodule measures 2 cm or more. These factors can increase false negatives to unacceptable levels. The 5 mm threshold is a useful benchmark, as sensitivity increases significantly for tumors above this size. Given the limitations in diagnosing multinodular goiters, stemming from unreliable FNA results, US false positives, and the typically slow growth of PTMCs, we recommend avoiding FNA in these cases and opting for direct surgical referral.

Using FNA in conjunction with ultrasound provides a clear diagnostic advantage, especially for subcentimeter tumors. Lastly, increased procedural experience significantly improved sensitivity, yielding a 23.5% gain throughout the study period, despite consistent tumor size and nodularity characteristics. This highlights the importance of institutional learning in maximizing cytological accuracy for these challenging tumors. We recommend using a 5 mm cutoff as the standard for FNA performance.

Take-home messages from this retrospective study: (1) A positive result can be acted upon with high confidence, but a negative result cannot reliably exclude malignancy; (2) tumors smaller than 5 mm represent a diagnostic blind spot for FNA—fortunately, these are generally less clinically significant; and (3) while experience, procedural improvements, and good case selection yield meaningful gains in performance, even under the most optimistic scenario, one-third of malignancies remain undetected in a non-screening setting.

## Data Availability

All data produced in the present study are available upon reasonable request to the authors

## I acknowledge the following

1. The data that support the findings of this study are available on request from the corresponding author. The data are not publicly available due to privacy or ethical restrictions.
2. No funding received.
3. No conflict of interest.
4. Author’s contribution:

- Imad Abdien EL Hag - conceptualization, statistical analysis, and manuscript writing (lead).
- Mohamed T Al Shammary reported and revised the ultrasound findings (equal).
- Raghad Tallab-collected and categorized the preliminary data (equal).
- S Patil – writing (equal).
- Abrar G Alghamdi edited the manuscript (equal).

---

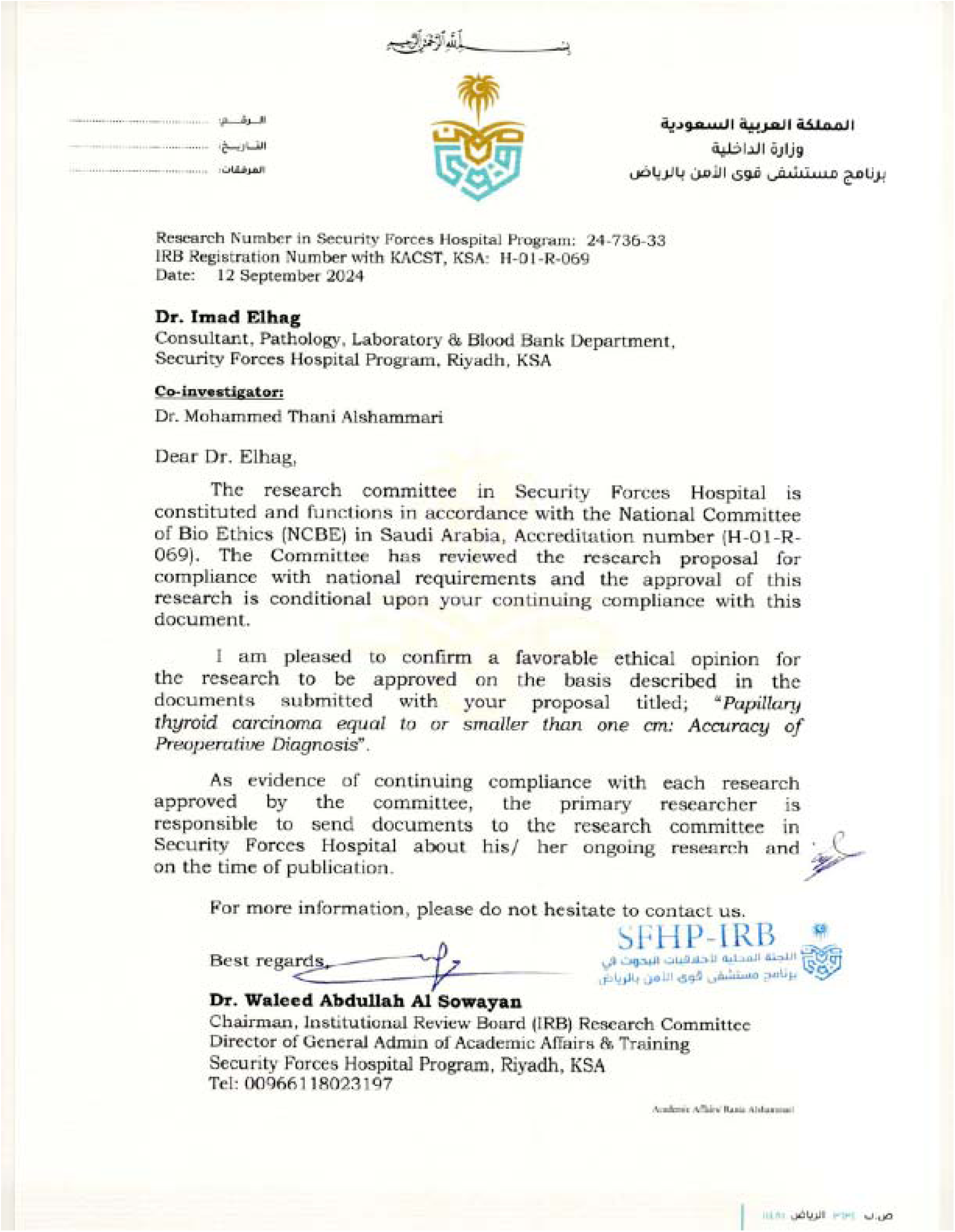

