## Supplementary material for "US-Guided Fine-Needle Aspiration of Papillary Thyroid Microcarcinoma in Non-Screening Settings: Diagnostic Errors and Mitigation Strategies": tables

Table 1. False negative rate in FNA across different tumor sizes.

| Variable | FN | TP | FN rate | OR (95% IC) | P vales |
| --- | --- | --- | --- | --- | --- |
| Group A (1- 10 mm) | 100 | 80 | 55.6 % | 9.64 (4.34 – 21.4) | < 0.001 |
| Group B (≥ 5 mm) | 45 | 71 | 39.0 % |  |  |
| Group C (< 5 mm) | 55 | 09 | 85.9% |  |  |

Table 2. Impact of DNs included vs. excluded on FNA results

| Variable | DNs included | DNs excluded | Total |
| --- | --- | --- | --- |
| TP | 72 (67.9%) | 8 (10.8%) | 69 (38.3%) |
| FN | 34 (32.1%) | 66 (89.2%) | 111 (61.7%) |
| Total | 106 | 74 | 180 |

Table 3. False negative rate in US across different tumor sizes

| Variable | FN | TP | FN rate | OR (95%IC) | P vales |
| --- | --- | --- | --- | --- | --- |
| Group A (1 -1 0 mm) | 90 | 90 | 50% | 0.17 (0.089 –0.33) | <.0001 |
| Group B (≥ 5 mm) | 41 | 75 | 35.3% |  |  |
| Group C (< 5 mm | 48 | 15 | 76.2% |  |  |

Table 4. Impact of DNs included vs. excluded on US results

| Variable | Absence of DNs | Presence of DNs | Total |
| --- | --- | --- | --- |
| TP | 70 (66%) | 20 (27%) | 90 |
| FN | 36 (34%) | 54 (73%) | 90 |
| Total | 106 | 74 | 180 |

Table 5. Causes and rates of US false positives

| Variable | TN | Observed FP | No | Expected  FP | OR | X^2^ | 95% IC | P value |
| --- | --- | --- | --- | --- | --- | --- | --- | --- |
| Calcification | 3 | 32 | 35 | 17.5 | 10.67 | 22.4 | 3.27 – 34.83 | <.0001 |
| Hypoechoic | 26 | 35 | 61 | 30.5 | 1.35 | 1.05 | 0.81 – 2.24 | 0.305 |
| Taller than wider | 0 | 04 | 04 | 02 | 9.0 | 2.25 | 0.48 – 167.17 | 0.134 |
| Granulomas | 0 | 02 | 02 | 01 | 5.0 | 0.5 | 0.24 – 104.15 | 0.480 |
